# Estradiol Modulates Resting-State Connectivity in Perimenopausal Depression

**DOI:** 10.1101/2024.06.05.24306430

**Authors:** Megan Hynd, Kathryn Gibson, Melissa Walsh, Rachel Phillips, Julianna Prim, Tory Eisenlohr-Moul, Erin Walsh, Gabriel Dichter, Crystal Schiller

## Abstract

The perimenopausal transition is marked by an increased risk for affective dysregulation and major depressive disorder (MDD), with hormone replacement therapy using estradiol (E2) showing promise for alleviating symptoms of perimenopausal-onset MDD (PO-MDD). Although E2’s effectiveness is recognized, its mechanisms underlying mood symptom modulation remain to be fully elucidated. Building on previous research suggesting that E2 may influence mood by altering cortico-subcortical connectivity, this study investigates the effects of transdermal E2 on resting-state functional connectivity (rsFC) in perimenopausal women with and without PO-MDD, focusing on rsFC changes using seed regions within emotion and reward processing networks. In this pharmaco-fMRI study, sixteen participants with PO-MDD and eighteen controls underwent rsFC analysis before and after three weeks of transdermal E2 administration. Pre-E2 results showed that the PO-MDD group, compared to controls, exhibited increased connectivity between the right amygdala (seed) and medial prefrontal cortex and anterior cingulate cortex, and decreased connectivity with the supplementary motor area. Post-E2 results revealed several significant E2-induced changes in connectivity between the PO-MDD and control groups: PO-MDD showed increased connectivity between the right caudate nucleus (seed) and left insula, and decreased connectivity between the right putamen (seed) and left hippocampus, and the right amygdala (seed) and left ventromedial prefrontal cortex. Notably, changes in connectivity were predictive of symptom trajectory across anhedonia, depressive mood, somatic, and vasomotor domains in the PO-MDD group. These findings enrich our understanding of PO-MDD by highlighting distinct rsFC patterns characteristic of the disorder and their shifts in response to E2 treatment, suggesting potential neural mechanisms underlying E2’s mood-modulating effects.

## Introduction

Perimenopause, the transitional phase preceding menopause, is associated with increased vulnerability to depressive symptoms and the development of major depressive disorder in susceptible women^1^. The ovarian hormonal fluctuations that accompany perimenopause, particularly estradiol (E2), have been implicated in the pathogenesis of perimenopause-onset major depressive disorder (PO-MDD)^2^. During perimenopause, women experience more rapid and larger magnitude changes (higher peaks and lower troughs) in E2 levels on a day-to-day basis than during pre-menopausal menstrual cycles^3^. This erratic and extreme E2 fluctuation has profound effects on brain neurochemistry, structure, and function^4^ and may disrupt processes such as emotion regulation and reward processing, which are implicated in MDD. Brief estrogen administration (i.e., three weeks) rapidly alleviates depressive symptoms in perimenopausal women^5^, yet the neural mechanisms by which E2 alleviates MDD are unknown. The purpose of this study is to investigate the effects of E2 on resting state functional connectivity (rsFC) in those with PO-MDD to identify the neural mechanisms underlying the rapid antidepressant effects of E2.

Several brain regions, specifically the striatum and amygdala, are implicated in depression and highly susceptible to the effects of gonadal steroid fluctuations^6–9^. These regions possess a high density of E2 receptors^10^ and demonstrate changes in rsFC in response to natural and exogenous E2 variations^11^. The pre-ovulatory stage of the menstrual cycle, coinciding with peak estradiol levels^11^, is characterized by increased frontostriatal connectivity. Exogenous E2 administration in postmenopausal women, leads to increased covariance between the amygdala and prefrontal cortex, as well as increased connectivity between the striatum and thalamus, as demonstrated by PET studies examining cerebral glucose metabolism and dopaminergic receptor activity, respectively^8,12^. Additionally, the use of neurosteroids (such as pregnenolone and DHEA) and oral contraceptives have been shown to modulate the functional connectivity of the amygdala with the salience and default mode network^13,14^ These alterations in connectivity underscore the complexity of hormonal influences on brain regions involved in emotion and reward processing, both of which are integral to the pathophysiology of PO-MDD.

Disruptions in connectivity involving the striatum and amygdala may contribute to the development and exacerbation of PO-MDD^4^. Exogenous E2 administration, thought to alleviate PO-MDD by stabilizing erratic E2 levels^5^, may exert its clinical effects by correcting aberrant striatum and amygdala connectivity patterns^4^. Decreased frontoamygdalar connectivity has been consistently found in individuals with MDD^15,16^, likewise, MDD is also characterized by decreased caudate connectivity with the default mode network^17,18^. Neuroimaging research using hormone administration (i.e., E2 with or without progesterone) in perimenopause is largely lacking, but two studies demonstrated E2 treatment increased task-based activation in the striatum and amygdala^19,20^. Yet, despite its potential for advancing biomarker development and therapeutic strategies, our understanding the impact of exogenous E2 on intrinsic functional connectivity in PO-MDD remains limited.

Investigating functional connectivity patterns may help us better understand the neural mechanisms underlying the rapid antidepressant effects of E2 during perimenopause. To isolate the antidepressant effects of E2 from the widespread effects of E2 across the perimenopausal brain, we examined the effects of transdermal E2 administration on the rsFC of striatum and amygdala seeds in women with PO-MDD compared with euthymic perimenopausal women (“controls”). Participants in both groups underwent a three-week transdermal E2 administration regimen, functional magnetic resonance imaging (fMRI) scans before and after administration (pre-E2 and post-E2, respectively), and repeated assessments of mood and menopause symptoms. We predicted that E2 administration would lead to increased connectivity involving striatal and amygdala seed regions in the PO-MDD group, relative to the control group. As described above, these hypotheses are supported by functional connectivity findings from studies investigating exogenous hormone administration and natural hormone fluctuation, respectively. Furthermore, we conducted an exploratory analysis to determine if pre-E2 level of connectivity and connectivity changes observed during administration predict the trajectory of improvements in mood symptoms, including anhedonia and depressed mood following E2 administration.

## Methods

### Participants

Forty-three perimenopausal women – defined in this study to include any person with intact ovaries – were recruited via social media advertisements, community flyers, and mass email messages to the University community and hospital patients. Participants were classified into two groups: (1) individuals with PO-MDD (n = 20), and (2) individuals who were free of Axis I disorders throughout their lifetime (n = 23). For both groups, inclusion criteria were ages 44-55 and late-stage perimenopause characterized by ≥ 60 days of amenorrhea or 2 missed menstrual cycles as defined by the Stages of Reproductive Aging Workshop (Stage -1)^21^. Exclusion criteria were current medication use, BMI < 18 or > 30 kg/m2, pregnancy or breastfeeding, hormonal contraindications (i.e., cardiovascular disease or personal or family history of breast cancer), MRI contraindications, recurrent MDD or MDD not related to perimenopause. Sample characteristics are presented in Table 1. The PO-MDD group had 16 participants with usable resting state fMRI data and the control group had 18 participants with usable data. Participants were not included in the analysis if they had missing resting-state scans or their movement during the scan exceeded the upper threshold of the third interquartile range.

**Table 1.** Baseline and post-treatment characteristics for the PO-MDD and the Control group. The PO-MDD group exhibited statistically significant reductions (p ≤ 0.05 uncorrected) in all symptom domains from pre-E2 to post-E2.

| Variables | Pre-E2 |  |  |  | Post-E2 |  |  |  |
| --- | --- | --- | --- | --- | --- | --- | --- | --- |
|  | PO-MDD | Control | t-statistic | p-value | PO-MDD | Control | t-statistic | p-value |
| Age (years), mean (SD) | 49.88 (2.76) | 49.39 (2.51) | 0.76 | 0.45 |  |  |  |  |
| **Race, n (%)** |  |  |  |  |  |  |  |  |
| White | 12 (75) | 15 (83) |  |  |  |  |  |  |
| African American or Black | 3 (19) | 0 (0) |  |  |  |  |  |  |
| Multiracial | 1 (6) | 0 (0) |  |  |  |  |  |  |
| Prefer not to answer/unknown | 0 (0) | 3 (17) |  |  |  |  |  |  |
| **Ethnicity, n (%)** |  |  |  |  |  |  |  |  |
| Hispanic or Latino | 1 (6) | 0 (0) |  |  |  |  |  |  |
| Not Hispanic or Latino | 15 (94) | 14 (78) |  |  |  |  |  |  |
| Prefer not to answer/unknown | 0 (0) | 4 (22) |  |  |  |  |  |  |
| **Marital Status, n (%)** |  |  |  |  |  |  |  |  |
| Married | 12 (75) | 13 (72) |  |  |  |  |  |  |
| Separated | 1 (6) | 1 (6) |  |  |  |  |  |  |
| Divorced | 1 (6) | 1 (6) |  |  |  |  |  |  |
| Single, never married | 2 (13) | 0 |  |  |  |  |  |  |
| Prefer not to answer/unknown | 0 | 3 (17) |  |  |  |  |  |  |
| **MASQ-AD, mean (SD)** | 67.19 (14.15) | 44.00 (8.90) | 5.64 | <0.001*** | 58.25 (17.07) | 43.11 (10.55) | 3.06 | .005** |
| **GCS, mean (SD)** |  |  |  |  |  |  |  |  |
| *Psychological* | 11.94 (5.72) | 2.22 (2.18) | 6.39 | <0.001*** | 5.75 (4.77) | 1.94 (2.53) | 2.85 | 0.01* |
| *Anxiety* | 5.50 (2.97) | 0.89 (0.96) | 5.95 | <0.001*** | 2.69 (2.15) | 1.00 (1.28) | 2.73 | 0.01* |
| *Depression* | 6.44 (3.88) | 1.33 (1.50) | 4.94 | <0.001*** | 3.06 (3.32) | 0.94 (1.59) | 2.33 | 0.03* |
| *Somatic* | 2.94 (3.17) | 0.72 (1.02) | 2.67 | 0.02* | 0.81 (0.83) | 0.72 (1.27) | 0.25 | 0.81 |
| *Vasomotor* | 2.56 (1.71) | 1.67 (1.28) | 1.71 | 0.10 | 0.38 (0.62) | 0.33 (0.59) | 0.20 | 0.84 |

### Procedures

#### Hormone Administration and Assessment

All participants received three weeks of transdermal E2 (100 ug/day via Climara® patch)^22^ following their pre-administration MRI session. Participants received an additional week of combined transdermal E2 (100 ug/day via Climara® patch) and micronized progesterone (200mg/day via Prometrium®)^23^ at the end of the study to precipitate menstruation and reduce the risk of endometrial hyperplasia. Plasma E2 and progesterone levels were collected at each imaging session and weekly study visits (see Figure 1).

**Figure 1.**
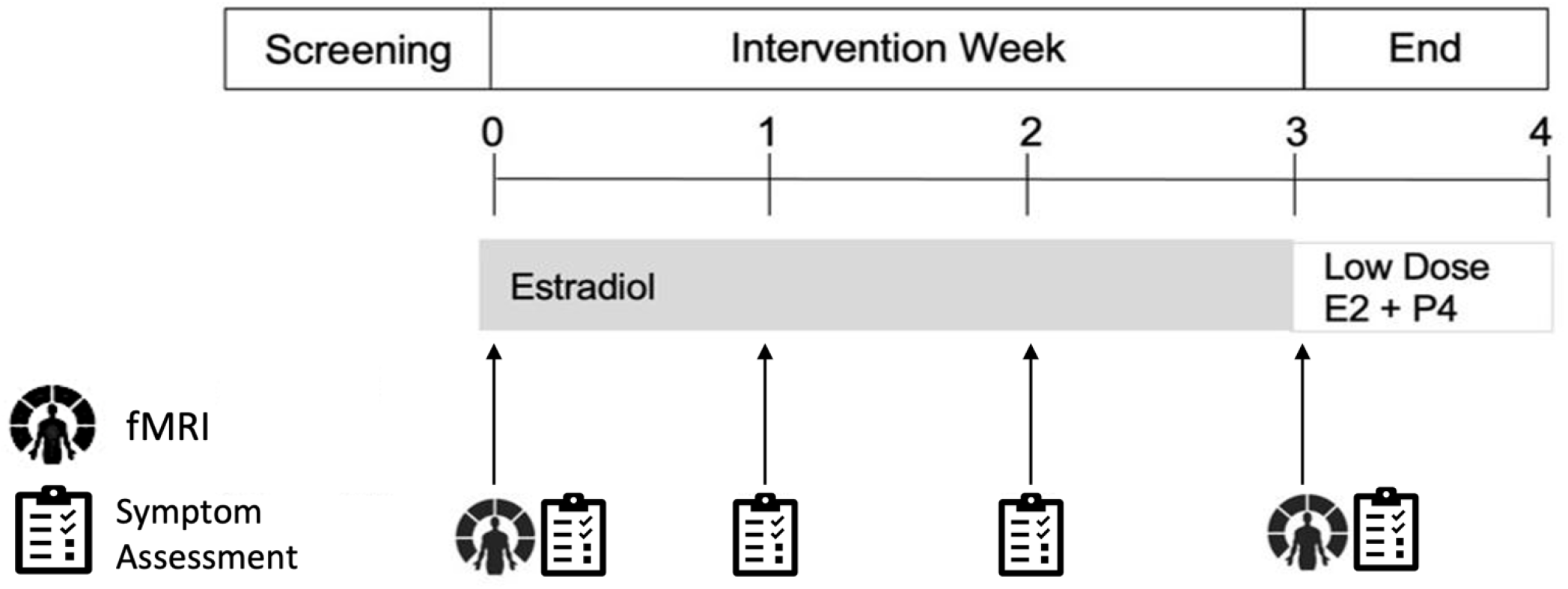
Treatment protocol. Imaging was acquired at pre-E2 (week 0) and post-E2 (week 3).

#### Clinical Assessments

Psychiatric illness was assessed via the Structured Clinical Interview for DSM-IV-TR for Axis I Disorders (SCID-IV)21^24^ at the baseline assessment to determine eligibility. The following symptom scales were administered at the baseline eligibility assessment, at baseline and follow-up MRI visits, and during weekly study visits: The Mood and Anxiety Symptom Questionnaire – Anhedonic Depression Subscale (MASQ-AD) is a 22-item scale that assesses anhedonia^25^; MASQ-AD scores have been associated with blunted striatal response to reward in previous studies of patients with depression^26^. The Greene Climacteric Scale (GCS) is a 21-item scale that assesses vasomotor, somatic, anxiety, and depressive symptoms of menopause^27^.

#### Imaging acquisition

Scans were acquired at the University of North Carolina at Chapel Hill using a 3T MAGNETOM TIM Trio Scanner. T1-weighted images used these parameters: echo time (TE) = 3.16 ms, repetition time (TR) = 2400 ms, inversion time (TI) = 1200 ms, flip angle = 8°, and voxel size of 1 × 1 × 1 mm. Resting-state data were acquired during a 5-minute (150 vol) gradient-echo echoplanar imaging sequence with the following parameters: TE = 32 ms, TR = 2000 ms, flip angle = 80°, and voxel size of 4.0 × 4.0 × 4.0 mm3, 36 axial slices. Participants were instructed to keep their gaze fixed on a crosshair and not to fall asleep.

#### Imaging analysis

Functional connectivity data were analyzed using CONN^28^ (RRID:SCR_009550) release 21.a^29^ and SPM^29^ (RRID:SCR_007037) release 12.7771.

Preprocessing: Functional and anatomical data were preprocessed using a preprocessing pipeline^30^ including realignment with correction of susceptibility distortion interactions, slice timing correction, outlier detection, direct segmentation, and MNI-space normalization, and smoothing. Functional data were realigned using SPM realign & unwarp procedure^31^, where all scans were coregistered to a reference image (first scan of the first session) using a least squares approach and a 6 parameter (rigid body) transformation^32^, and resampled using b-spline interpolation to correct for motion and magnetic susceptibility interactions. Temporal misalignment between different slices of the functional data (acquired in interleaved Siemens order) was corrected following SPM slice-timing correction (STC) procedure^33,34^, using sinc temporal interpolation to resample each slice BOLD timeseries to a common mid-acquisition time. Potential outlier scans were identified using ART^35^ as acquisitions with framewise displacement above 0.5 mm or global BOLD signal changes above 3 standard deviations^36,37^, and a reference BOLD image was computed for each subject by averaging all scans excluding outliers. Functional and anatomical data were normalized into standard MNI space, segmented into grey matter, white matter, and CSF tissue classes, and resampled to 2 mm isotropic voxels following a direct normalization procedure^38^ using SPM unified segmentation and normalization algorithm^39,40^ with the default IXI-549 tissue probability map template. Last, functional data were smoothed using spatial convolution with a Gaussian kernel of 6 mm full width half maximum (FWHM).

##### Denoising

Functional data were denoised using a standard denoising pipeline^41^ including the regression of potential confounding effects characterized by white matter timeseries (5 CompCor noise components), CSF timeseries (5 CompCor noise components), motion parameters and their first order derivatives (12 factors)^42^, outlier scans (below 28 factors)^36^, and linear trends (2 factors) within each functional run, followed by bandpass frequency filtering of the BOLD timeseries^41^ between 0.008 Hz and 0.09 Hz. CompCor^43,44^ noise components within white matter and CSF were estimated by computing the average BOLD signal as well as the largest principal components orthogonal to the BOLD average, motion parameters, and outlier scans within each subject’s eroded segmentation masks. From the number of noise terms included in this denoising strategy, the effective degrees of freedom of the BOLD signal after denoising were estimated to range from 68.2 to 80 (average 77.5) across all subjects^37^.

##### First-level analysis

Based on our hypotheses, we used a seed-to-voxel analysis with left and right seeds from the nucleus accumbens, caudate nucleus, putamen, and the amygdala. Seed-based connectivity (SBC) maps were estimated characterizing the patterns of functional connectivity with the 8 ROIs. Functional connectivity strength is represented by Fisher-transformed bivariate correlation coefficients from a weighted general linear model (weighted-GLM^45^), defined separately for each pair of seed and target areas, modeling the association between their BOLD signal timeseries.

Group-level analyses were performed using a General Linear Model (GLM^46^). For each individual voxel (for each SBC map) a separate GLM was estimated with group and time as independent variables and age used as a covariate. Voxel-level significance was evaluated using multivariate parametric statistics with random-effects across subjects and sample covariance estimation across multiple measurements. Inferences were performed at the level of individual clusters (groups of contiguous voxels) based on Gaussian Random Field theory^47,48^. Results were thresholded using a combination of a cluster-forming p < 0.001 voxel-level threshold, and a familywise corrected p-FDR < 0.05 cluster-size threshold per SBC map^49^.

### Planned analysis

Descriptive statistics were used to summarize the demographic and clinical characteristics of the participants, including age and clinical measures. Group differences in demographic and clinical variables were assessed using independent t-tests. Paired samples t-tests were used to assess symptom decrease from pre-E2 to post-E2 in the PO-MDD group. To investigate differences between the PO-MDD group and the control group with respect to the impact of three weeks of transdermal E2 administration on connectivity we examined Group (PO-MDD, control) × Time (pre-E2, post-E2). interactions on seed-to-voxel connectivity as noted above. Statistical thresholds were set at p < 0.01, corrected for multiple comparisons using FDR at p < 0.05. Significant interactions were probed with post hoc t-tests to investigate between and within group differences in connectivity.

To investigate how alterations in connectivity related to changes in symptom severity, we implemented a weighted multilevel model using self-reported symptom trajectories (four timepoints) as the dependent variable. A weighted multilevel model was used specifically to minimize the impact of multivariate outliers in analysis. Baseline connectivity and change in connectivity (follow-up – baseline) were used as the moderators of interest. Other fixed effects included in the models were visit number, baseline connectivity, the interaction of visit number and baseline connectivity, and the interaction of visit number and change in connectivity. A random intercept was included in the model to capture unexplained variability in participants’ self-report data not explained by fixed effects. We calculated the simple slope effects of each significant interaction (by using the ‘reghelper’ package^50^) to determine how different levels of connectivity at baseline and different levels of connectivity change during administration significantly influenced symptom trajectory.

## Results

### Resting-State Functional Connectivity (rsFC) Results

#### Pre-E2 Differences in seed-based rsFC between PO-MDD and Control groups

Pre-E2, the PO-MDD group showed significantly greater right amygdala (seed) connectivity with the medial prefrontal cortex (MNIxyz: -14 +58 +6, k=213, p<0.001) and the anterior cingulate cortex (MNIxyz: -10 +42 +12, k=86, p<0.001) compared with the control group. The PO-MDD group also exhibited significantly lower connectivity between the right amygdala (seed) and the supplementary motor area (MNIxyz: +8 -4 +56, k= 51, p<0.001) than the control group. No other group differences in seed-based rsFC emerged Pre-E2. See Figure 2 for more details.

**Figure 2.**
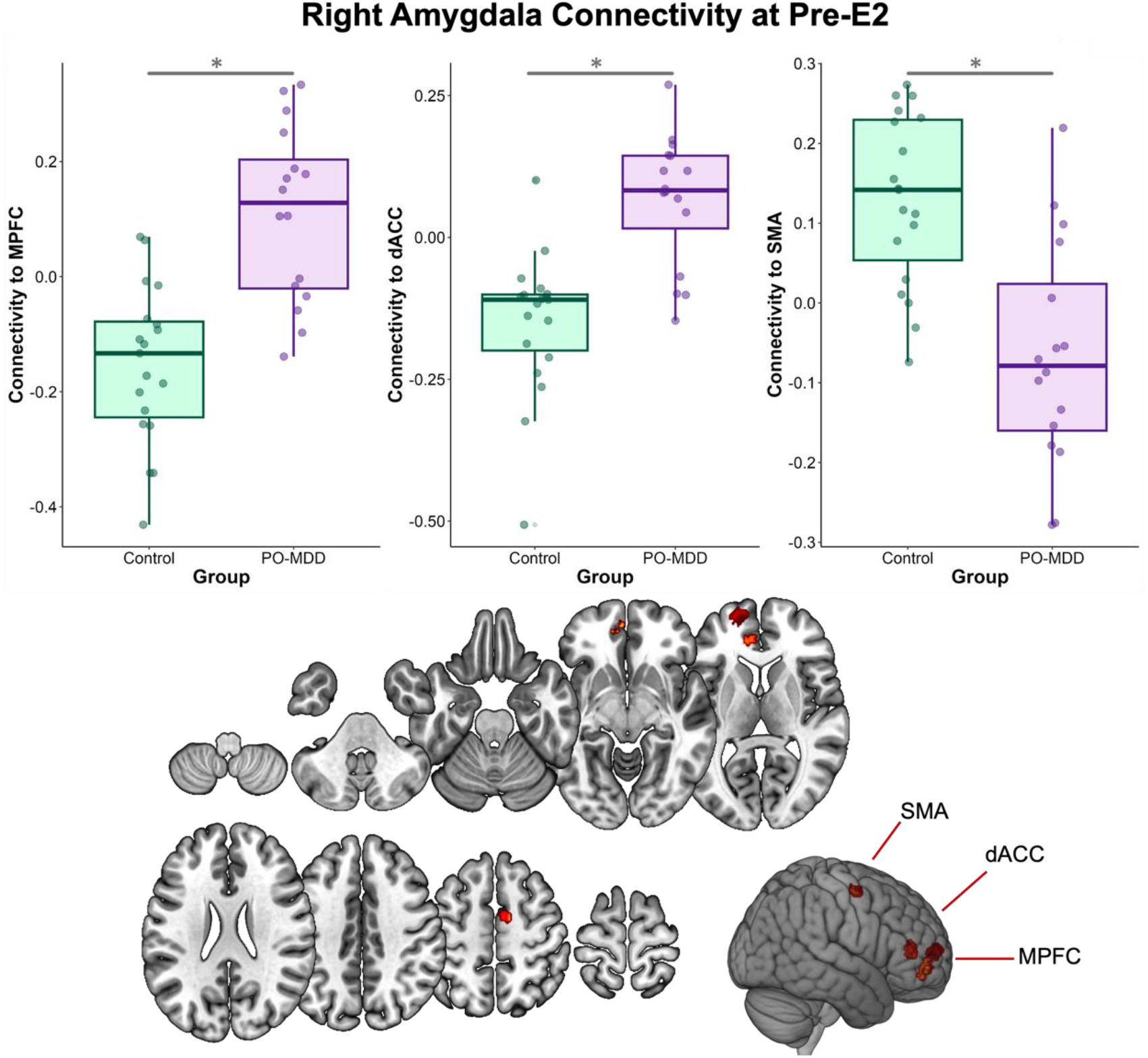
Baseline connectivity values using the right amygdala as the seed region. Connectivity values plotted based on group status. Target regions depicted on an MNI template. dACC – dorsal anterior cingulate cortex. MPFC – medial prefrontal cortex. SMA – supplementary motor area.

#### Effects of E2 on seed-based rsFC in PO-MDD and Control groups

We identified significant group × time interactions in the right caudate nucleus (seed) with the left insula (MNIxyz: -30 +18 +08, k=62, pFDR = 0.02), the right putamen (seed) with the left hippocampus (MNIxyz: -12 -44 +12, k=115, pFDR < 0.001), and the right amygdala (seed) with the left ventromedial prefrontal cortex (vmPFC; MNIxyz: -12 +50 -12, k=68, pFDR = 0.02), as shown in Figure 3. To explore whether changes in the PO-MDD or control group were driving significant findings, post-hoc analyses were conducted.

**Figure 3.**
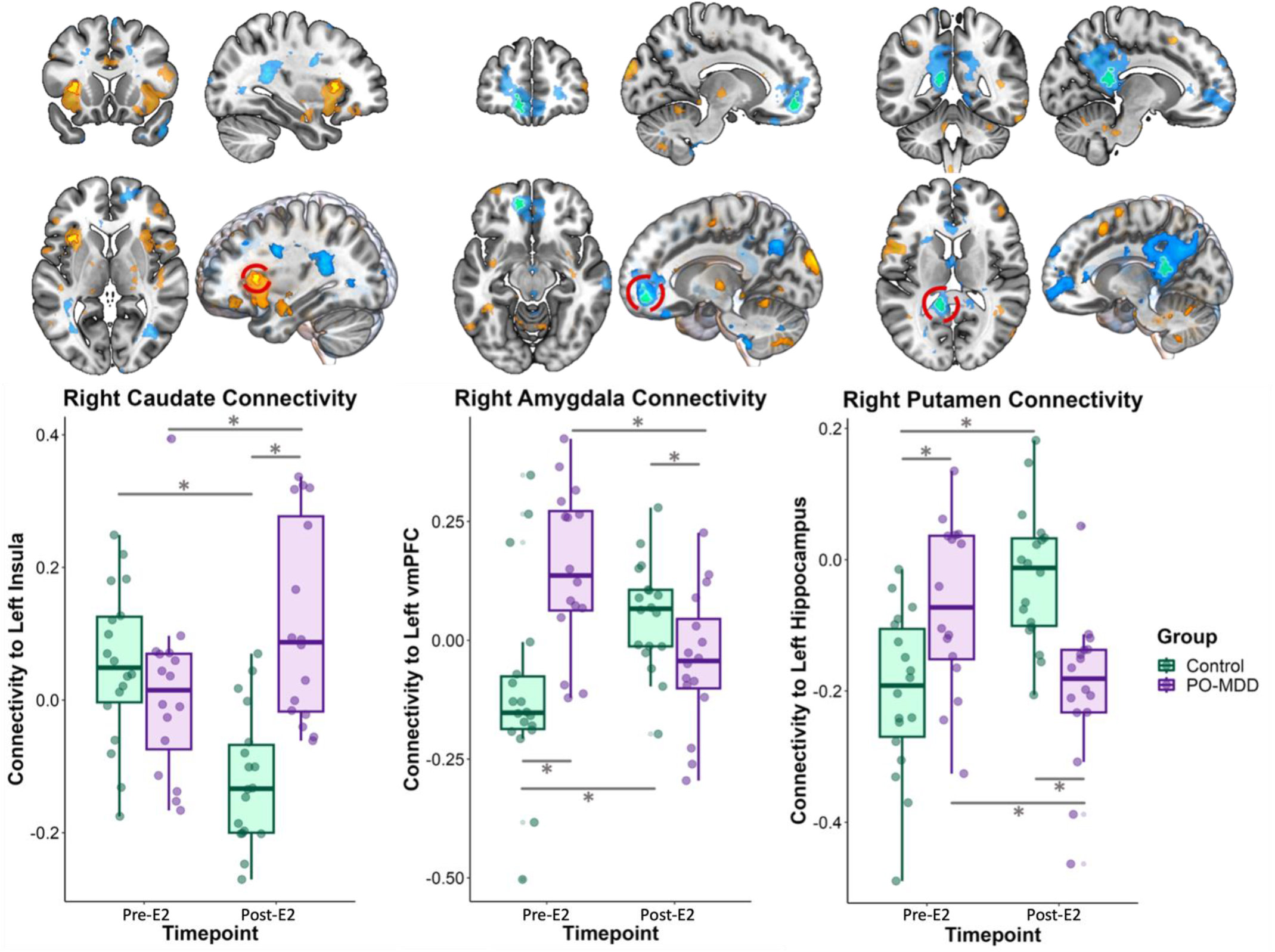
Differential effects of E2 on rsFC in PO-MDD and controls. Results shown are the significant connectivity findings from the group x time GLM analysis. The bottom panel shows individual connectivity at the pre-E2 and post-E2 time points separately for each group along with means and standard deviations. Above each plot is the target cluster displayed on an MNI template brain 3D render. The target cluster is outlined and circled in red. Outlined regions survived FDR correction, all other colored region scaled (darkest to brightest) from p<0.05 (uncorrected) ^53^. PO-MDD – perimenopausal-onset major depressive disorder. vmPFC – ventromedial prefrontal cortex.

A post-hoc independent samples t-test of caudate nucleus-insula connectivity at pre-E2 showed no significant differences in connectivity between groups. However, an independent samples t-test at post-E2 revealed a significant difference between groups, t(26) = 5.22, p<0.001, indicating that the PO-MDD group exhibited greater connectivity between the right caudate nucleus-left insula compared with the control group. Within-group analyses using paired t-tests demonstrated significant changes in connectivity from pre-E2 to post-E2 for both groups separately. Supporting our hypothesis, the PO-MDD group showed an increase in right caudate nucleus-left insula connectivity over time, t(15) = 2.51, p = 0.02, while the control group displayed a decrease in connectivity over the same period, t(17) = -4.6, p<0.001. Post-hoc independent samples t-tests showed that the PO-MDD group had significantly greater right putamen-left hippocampus connectivity at pre-E2, t(31) = 3.06, p = 0.005, but lower connectivity at post-E2, t(30) = -4.49, p < 0.001, compared with controls. Paired t-tests further indicated significant changes in putamen-hippocampus connectivity from pre-E2 to post-E2 in both groups. Contrary to our hypothesis, the PO-MDD group decreased connectivity, t(15) = -3.68, p = 0.002, while the control group increased connectivity, t(17) = 5.61, p < 0.001.

Post-hoc independent samples t-tests showed that the PO-MDD group had significantly greater right amygdala-left vmPFC connectivity at pre-E2, t(32) = 3.93, p<0.001, but lower connectivity at post-E2, t(28) = -2.12, p = 0.04, compared with controls. Paired t-tests further indicated significant changes in amygdala-left vmPFC connectivity from pre-E2 to post-E2 in both groups. Contrary to our hypothesis, the PO-MDD group exhibited a significant decrease in connectivity between the right amygdala and left vmPFC, t(15) = -4.10, p<0.001, while the control group demonstrated a significant increase in connectivity, t(17) = 2.80, p = 0.01).

### Resting-State Functional Connectivity as a Predictor of Clinical Outcomes in the PO-MDD Group

See Figure 4 for depiction of trajectory analysis using pre-E2 connectivity and connectivity change from pre-E2 to post-E2 as a moderator, respectively. Table 3 in the supplement shows simple slope analysis results. The relation between resting-state connectivity and clinical outcomes was only examined in the PO-MDD group.

**Figure 4.**
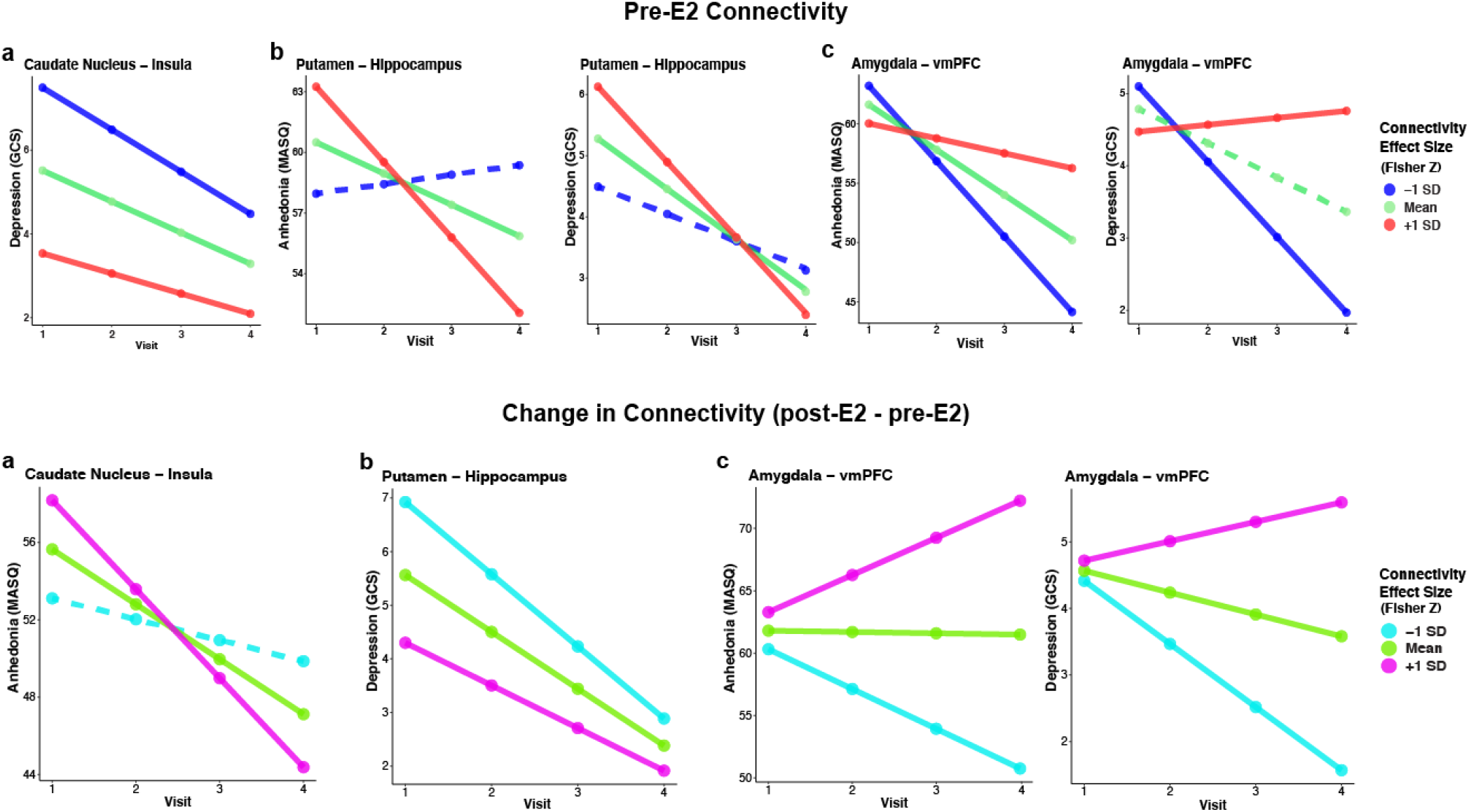
Trajectory Analysis of MASQ-AD and GCS Depression Subscales Across Four Timepoints, Moderated by pre-E2 (a) Caudate Nucleus-Left Insula Connectivity, (b) Right Putamen-Left Hippocampus Connectivity, and (c) Right Amygdala-Left vmPFC Connectivity displayed in the top panel and Moderated by connectivity change from pre-E2 to post-E2 displayed in the bottom panel. Solid lines statistically significant. See supplement for depiction of other GCS subscale trajectories.

#### Pre-E2 Connectivity Between the Right Caudate Nucleus and Left Insula as a Predictor of Symptom Trajectories Across E2 Administration

Within the PO-MDD group, right caudate nucleus-left insula connectivity at pre-E2 significantly predicted the subsequent trajectory of the GCS Depression subscale (p < 0.001), GCS Psychological subscale (p = 0.04), GCS Vasomotor subscale (p < 0.001), and GCS Somatic subscale (p < 0.001).

Specifically, in the GCS Depression and GCS Psychological subscale, lower pre-E2 connectivity predicted a consistently steeper decline in symptoms throughout the treatment course. Conversely, the GCS Vasomotor subscale demonstrated that higher pre-E2 connectivity was associated with a more pronounced reduction in vasomotor symptoms. Lastly, the GCS Somatic Symptoms subscale linked lower pre-E2 connectivity with a steeper decline in somatic symptoms, with significant negative trajectories.

#### Pre-E2 Connectivity Between the Right Putamen and Left Hippocampus as a Predictor of Symptom Trajectories Across E2 Administration

Within the PO-MDD group, right putamen-left hippocampus connectivity at pre-E2 significantly predicted the subsequent of the MASQ-AD (p < 0.001), GCS Depression subscale (p < 0.001), and GCS Psychological subscale (p = 0.03). Higher pre-E2 connectivity was associated with greatest decline in MASQ-AD anhedonia, GCS Depression and GCS psychological scale scores during treatment.

#### Pre-E2 Connectivity Between the Right Amygdala and Left vmPFC as a Predictor of Symptom Trajectories Across E2 Administration

Within the PO-MDD group, amygdala-vmPFC connectivity at pre-E2 significantly predicted symptom trajectories of the MASQ-AD (p < 0.001), GCS Psychological subscale (p = 0.03), GCS Depression subscale (p < 0.001), and GCS Vasomotor subscale (p = 0.01). Lower pre-E2 connectivity predicted a decrease in MASQ-AD symptoms, a greater reduction in GCS Depression symptoms, and a steeper decline in GCS Psychological symptoms. Conversely, higher pre-E2 connectivity predicted a more pronounced decrease in GCS Vasomotor symptoms.

#### Change in Connectivity Between the Right Caudate Nucleus and Left Insula as a Predictor of Symptom Trajectories Across E2 Administration

The change in right caudate nucleus-left insula connectivity with E2 administration significantly predicted the trajectory of symptom change in the MASQ-AD (p < 0.001), GCS Vasomotor subscale (p< 0.001), and GCS Somatic subscale (p < 0.001) in the PO-MDD group. Increased connectivity with E2 administration was associated with a more pronounced decrease in MASQ-AD anhedonia and GCS Vasomotor symptom scores. Conversely, decreased connectivity predicted a steeper downward trajectory of GCS Somatic symptom scores.

#### Change in Connectivity Between the Right Putamen and Left Hippocampus as a Predictor of Symptom Trajectories Across E2 Administration

The PO-MDD group’s change in right putamen-left hippocampus connectivity with E2 administration significantly predicted the trajectory of symptom change in the GCS Depression subscale (p < 0.001), the GCS Psychological subscale (p = 0.006), the GCS Anxiety subscale (p = 0.03), and the GCS Somatic symptom subscale (p < 0.001). Across all scales, decreased connectivity during E2 administration was associated with a steeper downward trajectory of symptoms.

#### Change in Connectivity Between the Right Amygdala and Left vmPFC as a Predictor of Symptom Trajectories Across E2 Administration

The change in right amygdala-left vmPFC connectivity with E2 administration significantly predicted the trajectory of symptom change in the MASQ-AD (p < 0.001) and GCS Depression subscale (p < 0.001). Decreased connectivity with E2 administration was associated with a more pronounced decrease in MASQ-AD anhedonia and GCS Depression scores.

## Discussion

This pharmaco-fMRI study examined the effect of E2 administration on resting-state functional connectivity in regions associated with emotion and reward in perimenopausal women with PO-MDD and those without MDD (i.e., “controls”). To our knowledge, this study is the first to examine the effect of E2 on connectivity in PO-MDD. Our findings provide evidence for possible neural mechanisms by which transdermal E2 modulates mood symptoms in PO-MDD. PO-MDD was characterized by greater connectivity from the right amygdala to the medial prefrontal and dorsal anterior cingulate cortex compared with the control group. Prior research on perimenopausal women and those with MDD have implicated connectivity in the same regions. Compared with premenopausal women, perimenopausal women show greater resting-state connectivity between the amygdala and medial prefrontal cortex and between the anterior cingulate and medial prefrontal cortex^51^. Further, hyperconnectivity between the medial prefrontal cortex and the amygdala have been identified as a potential biomarker of MDD^52^. Our results also showed that PO-MDD was characterized by less connectivity from the right amygdala to the supplementary motor area, which is largely involved in motor and sensory tasks^53^. This study did not explore the associations between baseline group differences in connectivity and clinical symptom trajectories, as it was primarily focused on the effects of estradiol on brain connectivity and associated changes in symptoms.

Our *a priori* hypotheses were generally supported: E2 administration differentially influenced striatal and amygdala connectivity in the PO-MDD group, and these alterations predicted symptoms reduction during the administration period. Though the hypothesized direction of effects – that E2 administration would increase connectivity from striatal and amygdala seed regions – was only supported by our caudate nucleus findings. E2 administration increased caudate nucleus-insula connectivity in PO-MDD, compared with the control group. Lower caudate-insula pre-E2 connectivity predicted faster reductions in clinical symptoms over the administration period while greater increases in connectivity from pre-E2 to post-E2 were associated with greater reductions in anhedonia and other depressive symptoms. The right caudate nucleus plays a crucial role in reward-prediction error, reward sensitivity, and emotion regulation^54^. Decreased caudate nucleus functional connectivity to the frontal lobes has been identified in prior studies of MDD, and greater alterations in connectivity is associated with greater severity of depressive symptoms^55^. Administration of estradiol and progestin (E2 + P4) have been shown to modulate caudate nucleus activity in perimenopausal women, demonstrating sex steroid regulation of this key reward processing region and suggesting a potential mechanism by which sex steroids modulate anhedonia^20^. The insula – which serves as a hub in the salience network^56^ – has shown aberrant connectivity to brain regions important for emotion and reward in MDD. For example, decreased functional connectivity between the insula and the DLPFC has been reported in fMRI studies of MDD^57,58^. Moreover, insula activity has been associated with hot flashes^59,60^ and sexual arousal^61^ in menopausal women. Thus, caudate-insula connectivity may play a central role in the antidepressant effects of E2 during perimenopause.

Our hypotheses regarding directionality were not supported by connectivity findings between the right putamen and left hippocampus. The PO-MDD group, compared with controls, showed decreased putamen-hippocampus connectivity during E2 administration. Those who started with higher putamen-hippocampus connectivity pre-E2 showed faster symptom reduction and those with the greatest decline in the connectivity with E2 administration showed the greatest decrease in anhedonia and other symptoms of depression. Thus, E2 may exert antidepressant effects by decreasing right putamen–left hippocampus connectivity in PO-MDD. The putamen is a critical structure within the basal ganglia, significantly influencing learning and reward sensitivity^62^. Hormone therapy (specifically E2 + P4) has been shown to enhance activity within the putamen during phases of reward anticipation, and putamen activity is correlated with endogenous E2 levels during reward acquisition^20^. Activation of the right putamen has also been shown to vary according to menstrual cycle phase^63^. The hippocampus is recognized for its role in learning, memory, spatial navigation, emotional behavior, and regulating hypothalamic functions^64^. Previous research has hormone replacement therapy (HRT) and E2 modulate hippocampus activity. In postmenopausal women, previous HRT use (E2 + P4) increases activity in the left hippocampus when compared with non-users^65^. Animal studies further reveal that E2 injections affect cholinergic pathways in the hippocampus, indicating possible shifts in neurotransmitter-related connectivity^66–69^.

Contrary to our directionality hypotheses, the PO-MDD group exhibited a significant decrease in right amygdala–left vmPFC connectivity during E2 administration, and those who showed lower connectivity pre-E2, as well as those with the greatest decrease in connectivity from pre-E2 to post-E2, experienced the greatest decrease in anhedonia and depression. The amygdala is fundamentally implicated in stimulus-reward learning and interacts with a myriad of brain regions, such as the striatum, basal forebrain, and the prefrontal cortex^70^. Notably, Berent-Spillson et al. (2017) and Ottowitz et al. (2008) emphasize the amygdala’s role in emotional processing and its variable activation in response to emotional states and estrogen levels, highlighting its central function in mood regulation. Moreover, the medial prefrontal cortex, particularly the vmPFC, significantly responds to emotionally negative stimuli and exhibits increased connectivity with the amygdala, underscoring its role in emotional regulation and mood disorders. Kaiser et al. (2015) further support this by showing major depression is associated with increased amygdala-medial prefrontal cortex connectivity, reinforcing the concept of a ‘medial prefrontal network’ crucial for mood regulation^71–73^. Our findings suggest E2 exerts antidepressant effects by decreasing amygdala– vmPFC connectivity in PO-MDD.

The discrepancy between our directional hypotheses and our findings may reflect underrepresentation of the hormonal basis of PO-MDD in current research. This condition, influenced by perimenopausal hormonal fluctuations, may elicit neural responses that differ from those typically seen in MDD. Furthermore, variations in research methodologies across studies, coupled with instability of results from small samples, may contribute to these discrepancies. Such findings underscore the need for more targeted research on this specific group with larger and methodologically consistent studies.

This study has limitations that should be considered when interpreting the results. First, there was no placebo control group, as all participants received estradiol. While the absence of a placebo control group hampers the ability to differentiate the effects of E2 from potential placebo effects, earlier studies demonstrated the antidepressant effects of E2 relative to placebo^5,74,75^. Second, in the context of this mechanistic trial, the post-E2 duration was relatively short, preventing the assessment of long-term outcomes beyond the study period. However, long-term studies of E2 efficacy have demonstrated maintenance of antidepressant effects for the duration of the treatment^76^.

Replication with a larger sample size, placebo-controlled design, and longer post-E2 period is necessary to determine the effects of E2 relative to placebo and to identify the long-term effects of E2 on brain function and connectivity. Functional connectivity patterns associated with other treatments commonly used in PO-MDD should also be investigated, including SSRIs and SNRIs. Moreover, prospective investigations on this topic could employ molecular imaging techniques to gain a deeper understanding of the molecular mechanisms associated with functional connectivity responses to E2 treatment. These future studies will contribute to a more comprehensive understanding of the underlying mechanisms and potential therapeutic implications of estradiol in the context of PO-MDD.

## Data Availability

All data produced in the present study are available upon reasonable request to the authors.

## Author contributions statement

M.H. led the conceptualization, formal analysis, and writing of the original draft. K.G., M.W., R.P., J.P., T.E.-M., and E.W. supported the formal analysis. G.D. provided supervision and supported the formal analysis. C.S. led the methodology, supervision, and supported the formal analysis. All authors reviewed and edited the manuscript.

