## Supplement for "Estradiol Modulates Resting-State Connectivity in Perimenopausal Depression"

**Supplemental Material**

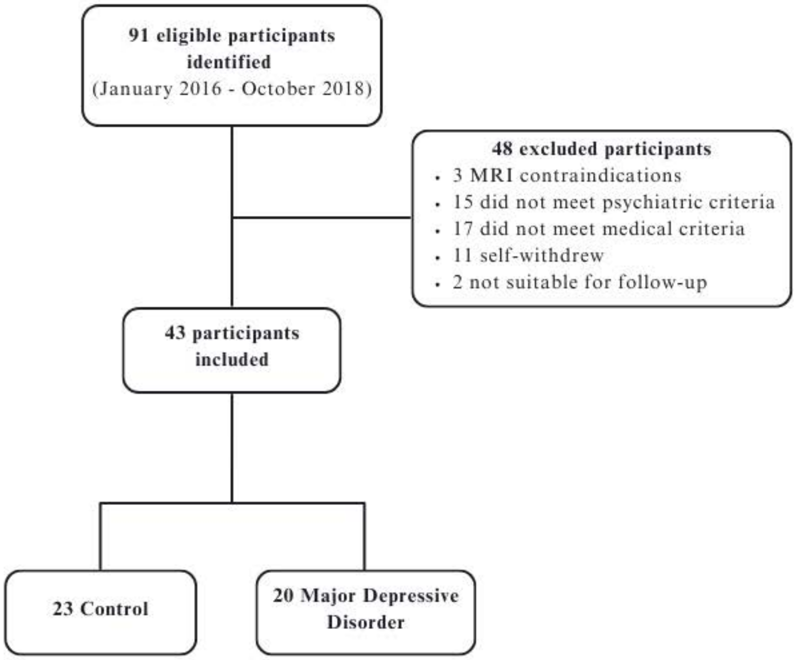


**Figure 4.** Study Consort Diagram.

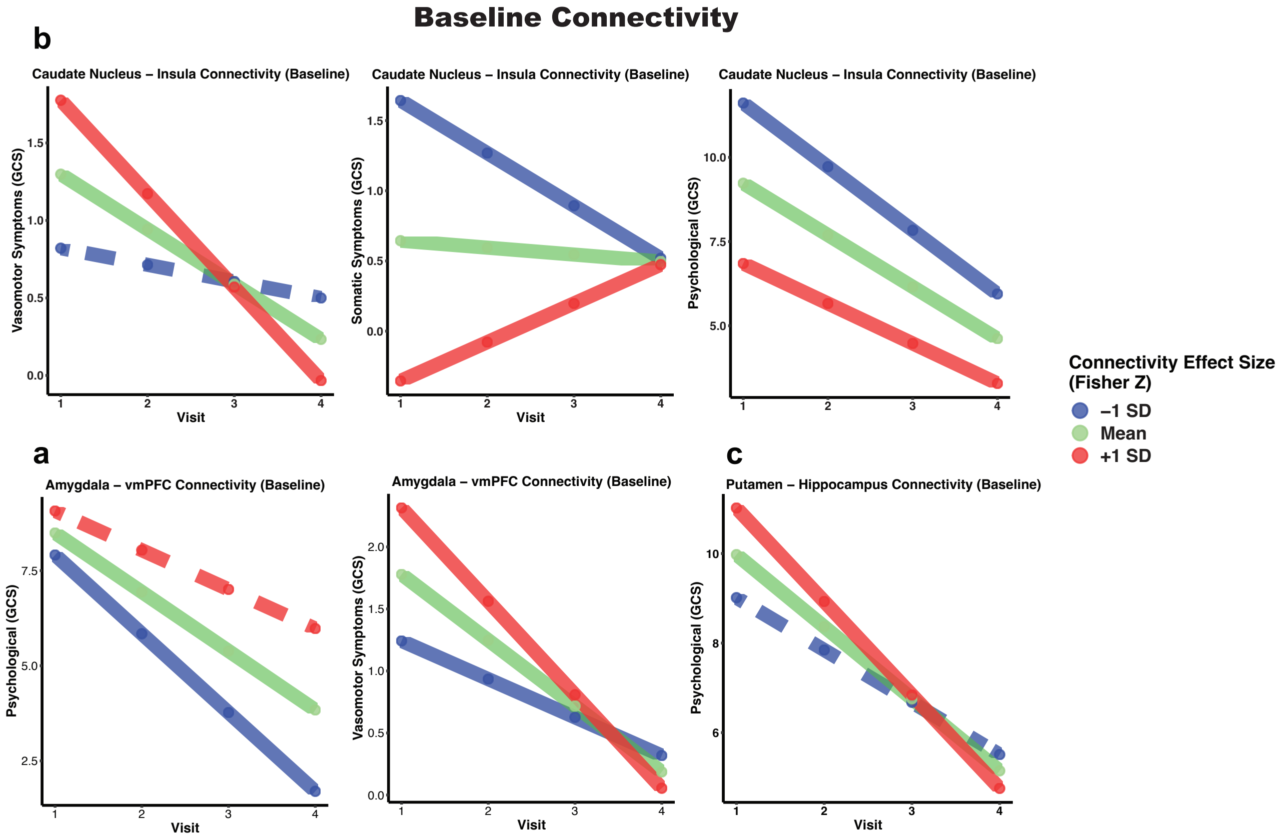


**Figure 5.** Trajectory Analysis of GCS Subscales Across Four Timepoints, Moderated by Baseline (a) Right Amygdala-Left vmPFC Connectivity, (b) Caudate Nucleus-Left Insula Connectivity, and (c) Right Putamen-Left Hippocampus Connectivity. Solid lines statistically significant.

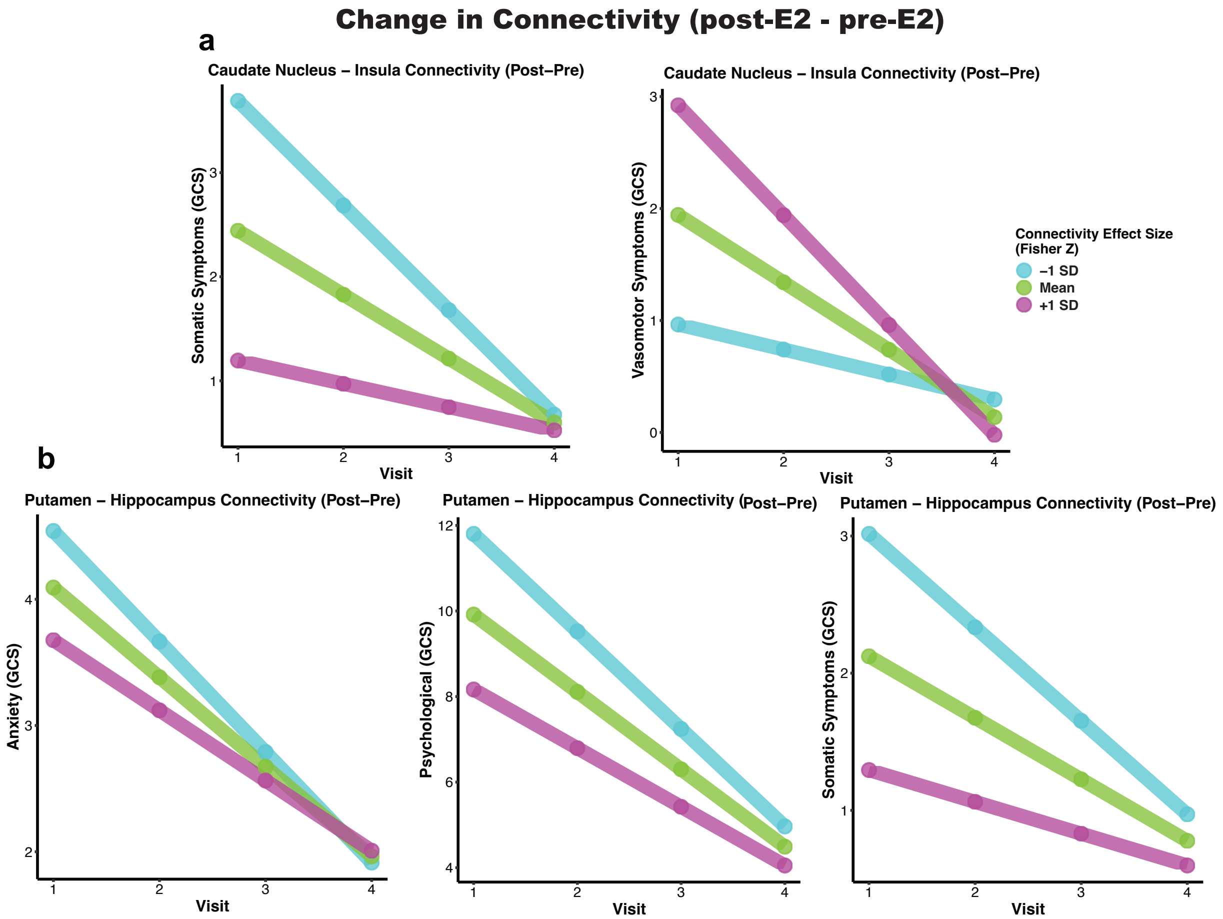


**Figure 6.** Predictive Analysis of Symptom Trajectory Over Treatment in GCS Subscales, Moderated by Change in (a) Right Caudate Nucleus-Left Insula Connectivity, (b) Right Putamen-Left Hippocampus Connectivity. Solid lines statistically significant.

| **A.** | **ROI Seed** | **Target (x, y, z)** | **Brodman Area** | **Target Name** | **Side** | **Target Cluster Size** | **GLM *p*-FDR** | **PO-MDD *t*-statistic (baseline)** |
|---|---|---|---|---|---|---|---|---|
| | **Right Amygdala** | | | | | | | |
| | | -14 +58 +6 | - | medial prefrontal cortex | L | 213 | < 0.001*** | 5.34 |
| | | -10 +42 +12 | 32 | anterior cingulate cortex | L | 87 | < 0.001*** | 5.31 |
| | | 8 -4 +56 | 6 | supplementary motor area | R | 51 | 0.048* | -4.54 |

| **B.** | **ROI Seed** | **Target (x, y, z)** | **Brodman Area** | **Target Name** | **Side** | **Target Cluster Size** | **GLM F-test statistic** | **GLM *p*-FDR** | **PO-MDD *t*-statistic (baseline vs follow-up)** |
|---|---|---|---|---|---|---|---|---|---|
| | **Right Amygdala** | | | | | | | | |
| | | 26 | 32 | ventromedial prefrontal cortex | L | 67 | 10.53 | 0.016* | 4.1 |
| | **Right Caudate** | | | | | | | | |
| | | -4 | - | insula | L | 59 | 15.17 | 0.021* | -2.51 |
| | **Right Putamen** | | | | | | | | |
| | | -44 | - | hippocampus | L | 115 | 21.14 | < 0.001*** | 3.68 |

**Table 2.** A) Significant baseline group differences in Baseline connectivity significant results using the right amygdala as the seed regionfrom a priori seeds. B) All Ssignificant group x time effects in connectivity results from a priori seeds, follow-up compared with baseline. GLM – general linear model. FDR – false discovery rate.

| | | **Right Caudate Nucleus and Left Insula** | | | **Right Amygdala and Left vmPFC** | | | **Right Putamen and Left Hippocampus** | | |
|---|---|---|---|---|---|---|---|---|---|---|
| | | -1 SD β*(SE)* | Mean β*(SE)* | +1 SD β*(SE)* | -1 SD β*(SE)* | Mean β*(SE)* | +1 SD β*(SE)* | -1 SD β*(SE)* | Mean β*(SE)* | +1 SD β*(SE)* |
| | **Symptom Measures** | | | | | | | | | |
| **Baseline Connectivity** | MASQ-AD | | | | -0.8(0.4) | 1.7(0.6) | **4.2(1.0)** | | -1.7(0.7) | **-3.8(0.9)** |
| | GCS *Psychological* | **-1.8(0.3)** | **-1.5(0.2)** | **-1.1(0.2)** | **-1.6(0.3)** | -1.1(0.4) | | | **-0.9(0.2)** | **-1.4(0.3)** |
| | GCS *Depression* | **-1.0(0.1)** | **-0.7(0.1)** | **-0.5(0.1)** | -0.6(0.2) | | 0.6(0.3) | | **-0.5(0.1)** | **-0.9(0.2)** |
| | GCS *Somatic* | **-1.1(0.1)** | **-0.8(0.1)** | **-0.5(0.1)** | | | | | | |
| | GCS *Vasomotor* | | **-0.4(0.1** | **-0.6(0.1)** | **-0.2(0.1)** | **-0.5(0.1)** | **-0.7(0.2)** | | | |
| | GCS *Anxiety* | | | | | | | | | |
| **Change in Connectivity** | MASQ-AD | | **-1.3(0.4)** | **-3.1(0.4)** | **-6.9(0.8)** | **-3.8(0.4)** | -0.7(0.4) | | | |
| | GCS *Psychological* | | | | | | | **-2.1(0.2)** | **-1.6(0.2)** | **-1.2(0.3)** |
| | GCS *Depression* | | | | -1.8(0.2) | **-1.2(0.2)** | -0.5(0.2) | **-1.3(0.1)** | **-1.0(0.1)** | **-0.7(0.1)** |
| | GCS *Somatic* | **-0.1(0.1)** | -0.6(0.1) | -0.2(0.03) | | | | **-0.7(0.1)** | **-0.4(0.04)** | **-0.2(0.04)** |
| | GCS *Vasomotor* | **-0.2(0.1)** | **-0.6(0.1)** | **-0.10(0.10)** | | | | | | |
| | GCS *Anxiety* | | | | | | | **-0.9(0.1)** | **-0.7(0.03)** | **-0.6(0.1)** |

**Table 3.** Simple Slope Analysis Results. All data points significant at uncorrected p < 0.05; bolded data points are significant at uncorrected p < 0.001.
